# Heteroresistance to first-line β-lactams among Gram-negative bloodstream isolates reported susceptible by standard-of-care testing: a prospective two-centre study across a high- and a low-resistance setting

**DOI:** 10.64898/2026.09.07.26362422

**Authors:** Damiano Squitieri, Giulia Menchinelli, Benjamin Berinson, Nicole Degel-Brossmann, Brunella Posteraro, Holger Rohde, Maurizio Sanguinetti

## Abstract

**Background:** Antibiotic heteroresistance (HR) — a resistant subpopulation within a clonal isolate reported susceptible by routine testing — can drive unexplained treatment failure and on-treatment emergence of resistance, yet prospective prevalence data for first-line β-lactams in Gram-negative bloodstream infection are lacking. We assessed the prevalence and genomic basis of HR to piperacillin, piperacillin/tazobactam, or meropenem among bloodstream isolates reported susceptible by standard-of-care testing.

**Methods:** In this prospective two-centre prevalence study we screened consecutive monomicrobial Gram-negative positive blood cultures at the University Medical Center Hamburg-Eppendorf (UKE), Hamburg, Germany, and the Fondazione Policlinico Universitario A. Gemelli IRCCS (FPG), Rome, Italy. Enterobacterales and Pseudomonadales reported susceptible by standard-of-care AST were allocated to one investigated beta-lactam (piperacillin, piperacillin/tazobactam or meropenem) according to their routine susceptibility phenotype (Vitek2), and entered a two-tier phenotypic workflow: an agar filter at 0.5x the EUCAST breakpoint in triplicate, followed - if positive - by population analysis profiling (PAP) up to 4x the breakpoint. Heteroresistance was defined as a clonal resistant subpopulation at a frequency of at least 0.5×10^-6^ surviving in the 4x breakpoint zone. Prevalence was estimated with Wilson-score 95% CIs and compared between centres with Fisher’s exact test. Confirmed heteroresistant isolates and their paired susceptible bulk populations underwent short- and long-read whole-genome sequencing, and each resistant/susceptible pair was tested for tandem gene amplification, differential subclonal point variants and structural variation.

**Findings:** 279 isolates were screened (93 at UKE, 186 at Gemelli). The 0.5x breakpoint filter was positive in 22 of 93 isolates (23.7%) at UKE and 66 of 186 (35.5%) at Gemelli. PAP confirmed heteroresistance in 6 of 93 isolates at UKE (6.5%, 95% CI 3.0-13.4) and 27 of 186 at Gemelli (14.5%, 10.2-20.3), a pooled prevalence of 33 of 279 (11.8%, 8.5-16.1); the between-centre difference did not reach significance (Fisher’s exact test p=0.051). Heteroresistance was essentially confined to piperacillin/tazobactam: 32 of 150 isolates screened against piperacillin/tazobactam were heteroresistant (21.3%, 15.5-28.6), versus 1 of 71 for piperacillin (1.4%) and 0 of 58 for meropenem. *Escherichia coli* accounted for 17 of the 33 heteroresistant isolates. In the resistant/susceptible pairs sequenced to date a candidate mechanism was resolved in 18: 13 carried a tandem amplification of a beta-lactamase locus (copy-number ratio 2.4-33.0; amplified units 5-136 kb, most often plasmid-borne and centred on *blaTEM-1, blaCTX-M* or *blaOXA-1*), and 5 carried point variants in the *AmpC* circuit (*ampD, ampR, dacB/PBP4*) or in efflux components, three of them fixed loss- or gain-of-function changes and three subclonal.

**Interpretation:** Roughly one in eight Gram-negative bloodstream isolates called susceptible to a first-line beta-lactam harboured a resistant subpopulation, and the burden fell almost entirely on piperacillin/tazobactam - the agent most often used empirically in this setting. The dominant genomic substrate was unstable amplification of an existing beta-lactamase gene rather than acquisition of a new resistance determinant, which explains why the phenotype is reversible, invisible to AST and, in principle, quantifiable by copy-number assays at the point of care.

**Funding:** Part of these data were presented at ESCMID Global 2026, and D.S. received an ESCMID travel grant in support of that presentation.

**Research in context:** *Evidence before this study:* Beta-lactam heteroresistance in Gram-negative bacteria has been characterised mainly in reference collections and in single-species surveys, where unstable tandem amplification of resistance genes was identified as the principal mechanism and shown to be reversible and frequently missed by routine AST. HR has been linked to unexplained treatment failure, shown to cause discrepant susceptibility results, and — in a recent report — demonstrated to underlie within-patient conversion from susceptible to resistant during therapy. However, prospective, multicentre prevalence data for HR to the first-line β-lactams used empirically in Gram-negative bloodstream infection (piperacillin, piperacillin/tazobactam, meropenem), restricted to isolates that routine AST reports as susceptible, and paired with genome-resolved mechanisms, were lacking.

*Added value of this study:* To our knowledge this is the first prospective, multicentre study to quantify PAP-confirmed HR to first-line β-lactams specifically among Gram-negative bloodstream isolates categorised as susceptible by standard-of-care AST, and to pair that prevalence with complete (short-plus long-read) genome analysis of clonal resistant/susceptible pairs. We show that HR is common (6.45% at UKE, 14.97% at FPG; pooled 12.1%, 34/280), that it is almost entirely confined to piperacillin/tazobactam (33 of 34 HR isolates; 21.9% of piperacillin/tazobactam-screened isolates versus ≤1.4% for piperacillin and none for meropenem), and that among 33 evaluable clonal pairs a genomic lesion is identifiable in 42·4%, dominated by unstable tandem amplification of β-lactamase-bearing blocks (11/33; 3·5–36×), with de-repressing mutations in efflux and AmpC regulators accounting for the remainder. Two further contributions are methodological. First, nearly half the pairs (45·5%) are negative on both genomic arms, placing non-genetic heteroresistance on the same footing as amplification rather than treating it as an exception. Second, resequencing the same isolates reproduced every amplification call but not its magnitude (+20% to −55%), showing that amplicon copy number is a snapshot of subclone frequency rather than a strain attribute — a constraint that any copy-number-based diagnostic must accommodate. Resistant subpopulations carried no evident fitness cost.

*Implications of all the available evidence:* AST-susceptible Gram-negative bloodstream isolates not infrequently harbour resistant subpopulations that current diagnostics cannot see but that can expand under first-line β-lactam therapy. Together with evidence that pre-existing HR can drive on-treatment resistance, our findings argue for the development of rapid, target-specific molecular assays (e.g. copy-number digital PCR) to detect HR in the diagnostic laboratory — read as quantitative, condition-dependent measurements rather than as fixed strain attributes — and for a prospective clinical study to determine whether breakpoint-crossing HR predicts therapeutic failure in Gram-negative bloodstream infection.

## Introduction

Antibiotic heteroresistance (HR) is the coexistence, within a genetically clonal bacterial isolate, of a minority resistant subpopulation alongside a susceptible majority [1,2]. Because routine antimicrobial susceptibility testing (AST) reports a single categorical result, the resistant subpopulation is usually invisible: the isolate is called susceptible, yet a fraction of cells can survive and expand under antibiotic exposure. First recognised for vancomycin in *Staphylococcus aureus* and penicillin in *Streptococcus pneumoniae* [3,4], HR is now documented across essentially every clinically important species–drug combination, and its population prevalence is high — driven, in Gram-negatives, chiefly by unstable tandem amplification of resistance genes [1,5].

The clinical stakes are increasingly clear. HR has been linked to unexplained antibiotic treatment failure [6], it is a demonstrable cause of discrepant AST results [7], and it can be the substrate from which stable, breakpoint-crossing resistance emerges during therapy: a recent Lancet Microbe report showed that a shift in pre-existing HR explained the within-patient change from susceptible to resistant during treatment [8], and HR to beta-lactams in particular may often be a way-station on the road to full resistance [9]. In *Escherichia coli* bloodstream infection, a retrospective cohort found HR to be common, frequently misclassified by routine testing, and clinically consequential [10]. Mechanistically, gene copy number is remarkably plastic: copy-number flexibility lets populations climb to higher resistance under rising drug pressure and threatens even new beta-lactam agents [11,12].

Despite this, prospective prevalence data for HR to first-line beta-lactams — the piperacillin/piperacillin–tazobactam/carbapenem backbone of empirical Gram-negative therapy — among isolates that routine AST reports as susceptible are scarce. Most systematic surveys have concentrated on last-line agents (colistin, cefiderocol) or on Gram-positive glycopeptide HR [5,13], and the phenotype’s dynamic, transient nature makes it hard to capture reproducibly in the diagnostic laboratory [14]. Population analysis profiling (PAP) remains the reference method, but it is labour-intensive and rarely applied prospectively at scale [1].

Gram-negative bloodstream infections (GN-BSI) are a setting where undetected HR could translate directly into empirical-therapy failure, yet the true burden of HR among culture-confirmed, AST-susceptible bloodstream isolates is unknown. We therefore conducted a prospective, multicentre study at two European tertiary centres to (1) determine the prevalence of PAP-confirmed HR to piperacillin, piperacillin/tazobactam, or meropenem among monomicrobial Gram-negative positive blood cultures reported susceptible by standard-of-care AST; (2) characterise the phenotype of the resistant subpopulations relative to the bulk population tested in the routine workflow; and (3) define, by whole-genome sequencing of clonal resistant/susceptible pairs, the genomic mechanisms underlying the differential beta-lactam susceptibility.

## Methods

### Study design and setting

We conducted a prospective, multicentre prevalence study at two European tertiary university hospitals: the University Medical Center Hamburg-Eppendorf (UKE), Hamburg, Germany, and the Fondazione Policlinico Universitario A. Gemelli IRCCS (FPG), Rome, Italy. Consecutive monomicrobial Gram-negative positive blood cultures (mGN-PBC) were screened during the enrolment periods June–November 2025 (UKE) and December 2025–May 2026 (FPG). The screening and confirmation cascade is summarised in Figure 1. The study is reported in accordance with the STROBE statement for observational studies.

**Figure 1:**
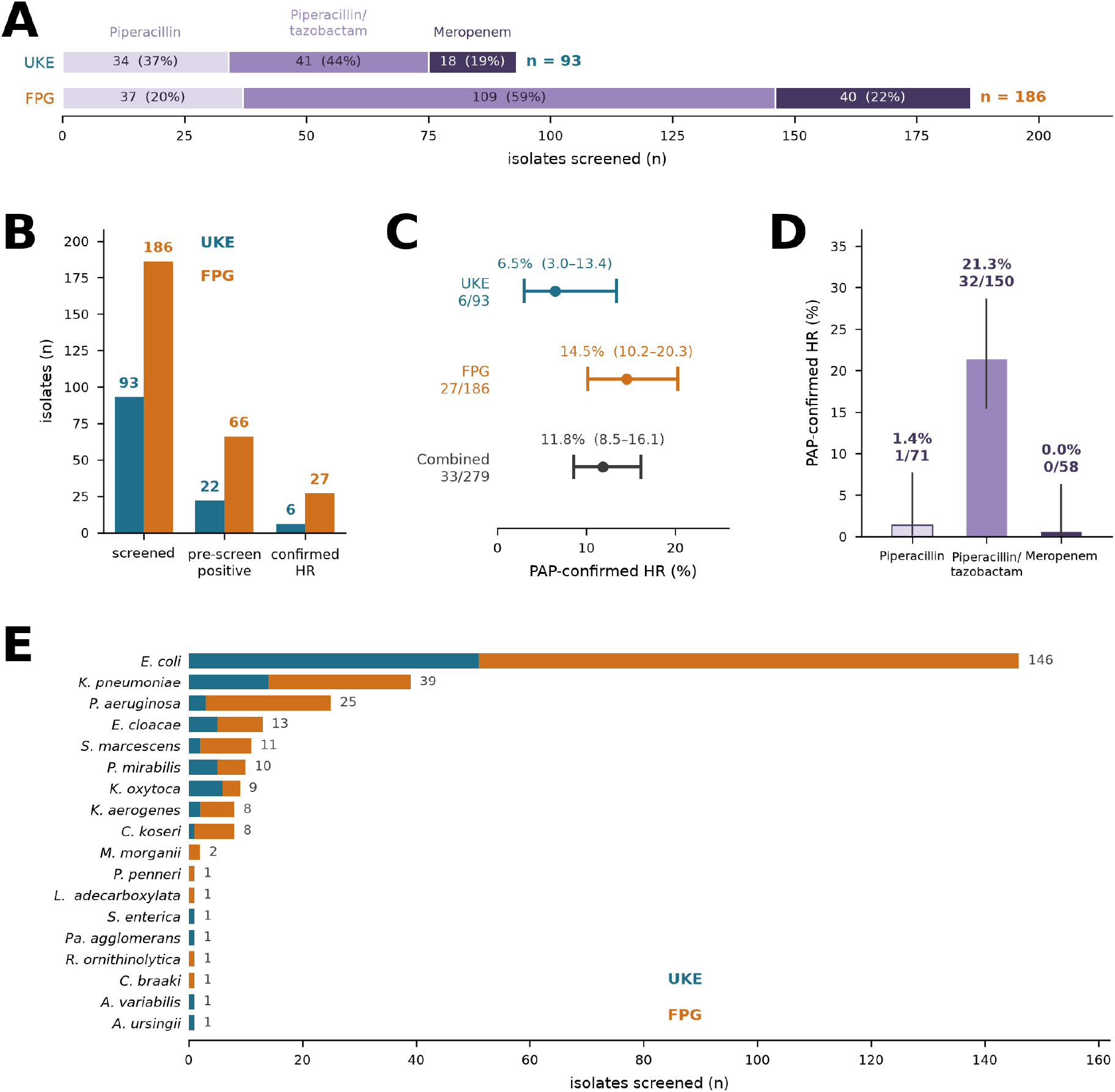
Enrolment, screening cascade and prevalence of heteroresistance. (A) Isolates screened at each centre, split by the investigated beta-lactam. (B) Screening cascade per centre: isolates screened, positive on the 0.5x breakpoint agar filter, and PAP-confirmed heteroresistant. (C) Prevalence of PAP-confirmed heteroresistance per centre and pooled, with Wilson-score 95% CIs. (D) Prevalence by investigated agent, with 95% CIs. (E) Species distribution of screened isolates by centre. In (B) and (E), blue denotes UKE and orange FPG (Fondazione Policlinico Universitario A. Gemelli IRCCS, abbreviated FPG in all figures); in (A) and (D), shading denotes the investigated agent (light, piperacillin; mid, piperacillin/tazobactam; dark, meropenem).

### Ethics

The study used only anonymised bacterial cultures recovered from routine positive blood cultures and involved no sensitive or identifiable patient data. Formal ethics-committee approval was therefore not required at either centre.

### Participants and eligibility

Eligible isolates were consecutive Enterobacterales and Pseudomonadales recovered from monomicrobial positive blood cultures, identified via MALDI-TOF (Bruker), and reported susceptible (S) to the investigated first-line β-lactam by standard-of-care AST (Vitek2, bioMérieux), interpreted per EUCAST clinical breakpoints v16.0 (2026). Polymicrobial cultures were excluded. For each isolate, the antibiotic taken forward to heteroresistance screening was assigned as follows: piperacillin if the isolate was S to piperacillin; piperacillin/tazobactam if S to piperacillin/tazobactam and R to piperacillin, or for *Pseudomonas* spp.; meropenem if R to piperacillin/tazobactam and S to meropenem, or for *Acinetobacter* spp. Pairs are pseudonymised as integers; no patient identifiers, admission dates or laboratory accession codes are used in any analysis or deposited file.

### Heteroresistance definition

Following the population-analysis framework established for β-lactam heteroresistance [1,5,9], an isolate reported susceptible by routine AST was classified breakpoint-crossing heteroresistant when a clonal resistant subpopulation, present at a frequency ≥0.5×10^−6^(i.e. above the spontaneous single-step mutation floor), survived on the population analysis profile (PAP) at antibiotic concentrations up to 4× the EUCAST breakpoint. Resistant subpopulations at a frequency ≥10^−4^ were designated high-frequency HR and those <10^−4^ low-frequency HR; because no standardised cut-off exists in the literature, this threshold is stated as a study convention.

### Two-tier phenotypic screening

Pre-screening (filter step). Three single colonies from the primary blood-culture subculture plate were inoculated into 1 mL Mueller–Hinton (MH) broth (Merck) and incubated for 18 h at 37 °C, 5% CO_2_, with shaking at 100 rpm. The suspension was plated in parallel onto antibiotic-free MH agar (growth control) and MH agar containing the investigated antibiotic at 0.5× the EUCAST breakpoint (BP/2) — an agar-dilution “microdilution-PAP” filter. The procedure was performed in triplicate from three separately picked single colonies. An isolate was pre-screening positive when at least one colony — confirmed by species identification to be the investigated organism — grew on the BP/2 plate in at least one of the three replicates.

Population analysis profiling (reference method). Pre-screening-positive isolates underwent full PAP. Standardised suspensions were plated by 10-fold serial dilution onto MH agar containing the investigated antibiotic at 0, 0.5, 1, 2 and 4× the EUCAST breakpoint. After incubation, colonies were counted and the surviving fraction at each concentration was calculated relative to the antibiotic-free control. Isolates meeting the heteroresistance definition (a resistant subpopulation surviving in the 2–4× breakpoint “HR zone”) were confirmed as heteroresistant, and the subpopulation frequency assigned the high- or low-frequency class. To confirm Vitek 2, broth microdilution MICs of piperacillin and piperacillin/tazobactam were determined for the bulk population and the PAP-recovered resistant subpopulation of each confirmed HR isolate.

### Whole-genome sequencing and genomic analysis

Thirty-three clonal resistant/susceptible (R/S) isolate PAP-positive pairs were investigated genomically. The 66 isolates were identified as follows: *E. coli* (n = 34), *Pseudomonas aeruginosa* (12), *Enterobacter cloacae* (8), *Klebsiella pneumoniae* (8), *Klebsiella aerogenes* (2) and *Citrobacter koseri* (2). Each pair comprises two isolates from the same episode in the same patient, differing in phenotype to the beta-lactam under study. Every comparison in this study is made within a pair. Within each pair, one arm’s assembly is the reference for all downstream stages, so that both arms are analysed in one coordinate frame. That arm is the susceptible isolate in 22 pairs and the resistant isolate in 11 (per-pair table in the repository); coordinates and reference-relative allele fractions are therefore interpretable within a pair and not across pairs. Confirmed HR isolates and their paired susceptible bulk populations underwent whole-genome sequencing combining short-read (DNBSEQ, BGI) and long-read (CycloneSEQ, BGI) chemistries, enabling complete, closed assemblies Each isolate was assembled from its long reads alone with Flye 2.9.6 (--nano-hq, nominal genome size 5 Mb) from a subsampled read set; assemblies comprise 1–3 contigs. Short reads were not used for assembly or polishing — they serve as an independent consistency check on the coverage-based dosage estimates below.

Genomes were annotated with Bakta v1.12.1 (database v6.0 light) and resistance determinants inventoried with AMRFinderPlus. The subsampling depth, the sequencing platform and library chemistry, read pre-processing, and the AMRFinderPlus and species-call database releases could not be recovered from the surviving working files and are stated as unrecovered rather than reconstructed (see Provenance below). Variants were called jointly for the two arms in a single two-sample pileup against the pair’s reference assembly (bcftools 1.24/htslib 1.24, mpileup -q 20 -Q 20 -a AD → call -mv → norm; command line recovered verbatim from the call-set headers). Joint two-sample calling is what makes an arm-versus-arm allele-fraction comparison meaningful at a site, and retained allelic depths are what make the classification below possible. Distances are taken from read evidence, not from assembly-versus-assembly comparison over sliding windows: in that approach paralogue misalignment within repeat families inflates mismatches by an amount that shifts with the window boundaries, so it is not correctable by a scaling factor. The superseded counts are not carried forward. Requiring ≥ 10 reads in both arms, sites were classed as fixed (one arm ≥ 0.90 non-reference fraction, the other ≤ 0.10), subclonal (≥ 0.20 vs ≤ 0.05, differing by ≥ 0.20) or shared — the last departing from the reference consensus without differing between arms, and excluded. Only the largest contig was treated as the chromosome; runs of ≥ 5 differential sites spaced ≤ 500 bp were collapsed to one event; indels were counted separately and never added to the substitution distance; and every discarded call is reported in audit columns, so a distance reconciles with its raw call set. The reported distance is dispersed fixed substitutions plus dense tracts, each tract counted once, and is 0–4 per pair (median 1) across the 17 pairs, with two pairs additionally carrying a tract. The classification uses the maximum and minimum of the two allele fractions and is therefore symmetric in the arms. Permissive subclonal counts are threshold-dominated — allele fractions pile up immediately above the 0.20 cut-off in depth-depleted arms — so only a filtered count (≥ 20 supporting reads, between-arm depth ratio ≥ 0.5) is reported. The 16 pairs without a call set receive no distance, and no value from the superseded method is substituted for them. All 66 genomes were compared pairwise by k-mer sketch (Mash v2.3, sketch -k 21 -s 200000, then dist). These parameters did not survive in any command line and were identified by re-executing the stage over a grid of 11 k-mer lengths × 7 sketch sizes against the archived distance matrix: k = 21 with sketch size 200 000 is the only combination that reproduces it, to the six significant digits at which the matrix was stored. Sketch distances are reported as such — not converted to SNP counts and not compared with published clonality thresholds, which address inter-patient transmission rather than two isolates from one episode. Missense variants were scored with ESM-2 650M (esm2_t33_650M_UR50D) as the masked-marginal log-likelihood ratio between mutant and wild-type residue, with relative solvent accessibility, per-residue pLDDT and distance to the catalytic site taken from AlphaFold DB models. The analysis was reconstructed from surviving intermediate files rather than a contemporaneous run log, and the repository separates parameters recovered verbatim from tool-written headers and logs. Unrecovered parameters are declared, not replaced by plausible defaults, and each is listed with the specific file that would close it.

### Data and code availability

Raw reads for all 101 libraries from the 66 isolates will be deposited in the European Nucleotide Archive under BioProject PRJEBXXXXXX (66 samples against checklist ERC000028; 66 long-read and 35 short-read runs). Sample and run metadata prepared for deposition, and the derived tables underlying every figure and table in this manuscript, are in the repository below, so deposited records and analysis inputs can be checked against each other. Before submission, replace PRJEBXXXXXX with the accession returned by Webin and add the sample accession range. No ENA accession exists for this study until deposition and none should be cited. Two items must be settled first: per-isolate collection dates, which the checklist requires and which survive in no analysis file, and a genus-level species discordance on one pair (*E. coli* by genomic call vs *C. koseri* in the clinical database). All analysis code, with the exact command lines, tool versions and thresholds used at every stage, and with the per-stage record of which parameters were recovered and which could not be, is public at https://github.com/damianosquitieri96-hr/hr-heteroresistance-pipeline (release v1.0.1, archived at Zenodo, doi:10.5281/zenodo.22288189).

### Fitness and copy-number quantification (planned analyses)

To assess whether the resistant subpopulations carry a fitness cost, growth-curve and time–kill assays comparing the resistant subpopulation with the paired bulk population are planned. As it stands, these data are available only for the UKE PAP-positive strains.

### Statistical analysis

Prevalence was expressed as a proportion with 95% Wilson-score confidence intervals, calculated per centre and pooled. Between-centre comparison of HR prevalence used Fisher’s exact test; categorical variables were compared with Fisher’s exact or χ^2^ tests, with a two-sided α of 0.05. Analyses were performed in Python 3 (scipy.stats).

## Results

### Enrolment and pre-screening

279 consecutive monomicrobial Gram-negative bloodstream isolates reported susceptible to a first-line beta-lactam by routine AST were screened: 93 at UKE and 186 at FPG (figure 1A, 1B). Allocation to the investigated agent differed between centres, reflecting the local susceptibility phenotype mix: piperacillin 34 of 93 (37%) at UKE and 37 of 186 (20%) at Gemelli, piperacillin/tazobactam 41 (44%) and 109 (59%), meropenem 18 (19%) and 40 (22%). *E. coli* was the predominant species at both centres (146 of 279 overall), followed by *K. pneumoniae* (39), *P. aeruginosa* (25) and *E. cloacae* complex (13), with *Serrati marcescens, Proteus mirabilis, Klebsiella oxytoca, K. aerogenes* and *C. koseri* each contributing 8-11 isolates (figure 1E).

The agar filter at 0.5x the EUCAST breakpoint was positive - at least one colony of the investigated organism on the antibiotic-containing plate in at least one of three replicates - in 22 of 93 isolates at UKE (23.7%, 95% CI 16.2-33.2) and 66 of 186 at Gemelli (35.5%, 29.0-42.6). These 88 isolates entered confirmatory PAP.

### Prevalence of PAP-confirmed heteroresistance

PAP confirmed a resistant subpopulation surviving in the 2-4x breakpoint zone in 6 of 93 isolates at UKE (6.5%, 95% CI 3.0-13.4) and 27 of 186 at Gemelli (14.5%, 10.2-20.3), giving a pooled prevalence of 33 of 279 (11.8%, 8.5-16.1) (figure 1C). The between-centre difference was of borderline significance (Fisher’s exact test, OR 0.41, p=0.051) and is compatible with the different allocation mix, since Gemelli screened a substantially larger share of its isolates against piperacillin/tazobactam. Heteroresistance was almost entirely a piperacillin/tazobactam phenomenon (figure 1D). Among isolates screened against piperacillin/tazobactam, 32 of 150 were heteroresistant (21.3%, 95% CI 15.5-28.6), compared with 1 of 71 screened against piperacillin (1.4%, 0.2-7.6) and 0 of 58 screened against meropenem (0%, 0-6.2). All 27 heteroresistant isolates at FPG and 5 of the 6 at UKE were piperacillin/tazobactam-heteroresistant; the single piperacillin case was a *K. aerogenes* isolate at UKE. Of the 33 heteroresistant isolates, 17 were *E. coli*, 6 *P. aeruginosa*, 4 *E. cloacae* complex, 4 *K. pneumoniae*, and one each *K. aerogenes* and *C. koseri*.

### Genomic mechanisms

Every sequenced resistant/susceptible pair was clonal (average nucleotide identity at least 99.99%, a handful of genome-wide SNPs and indels; figure 3A-3D), so the resistant population arose within the patient isolate rather than from co-infection. Mechanism assignment by centre and by species is shown in figure 2A and 2B.

**Figure 2:**
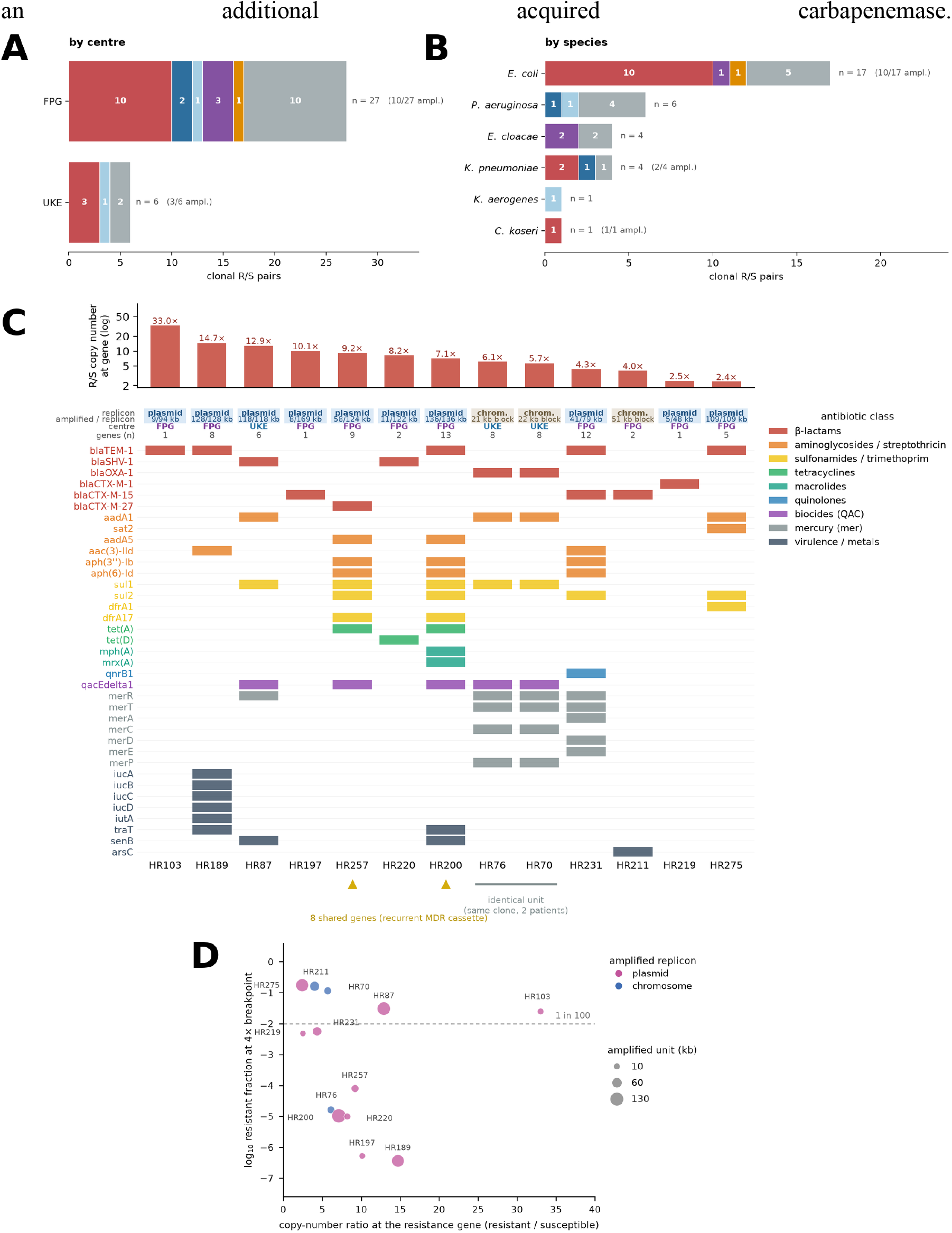
Genomic mechanisms of heteroresistance. (A) Mechanism assigned to each resistant/susceptible pair, by centre. (B) The same assignment by species. (C) Gene content of the amplified units, one row per pair. (D) Amplified unit length against copy-number ratio; symbol area scales with the number of resistance genes inside the unit and colour denotes the replicon (blue, chromosome; magenta, plasmid). The dashed line marks a copy-number ratio of 1.

**Figure 3:**
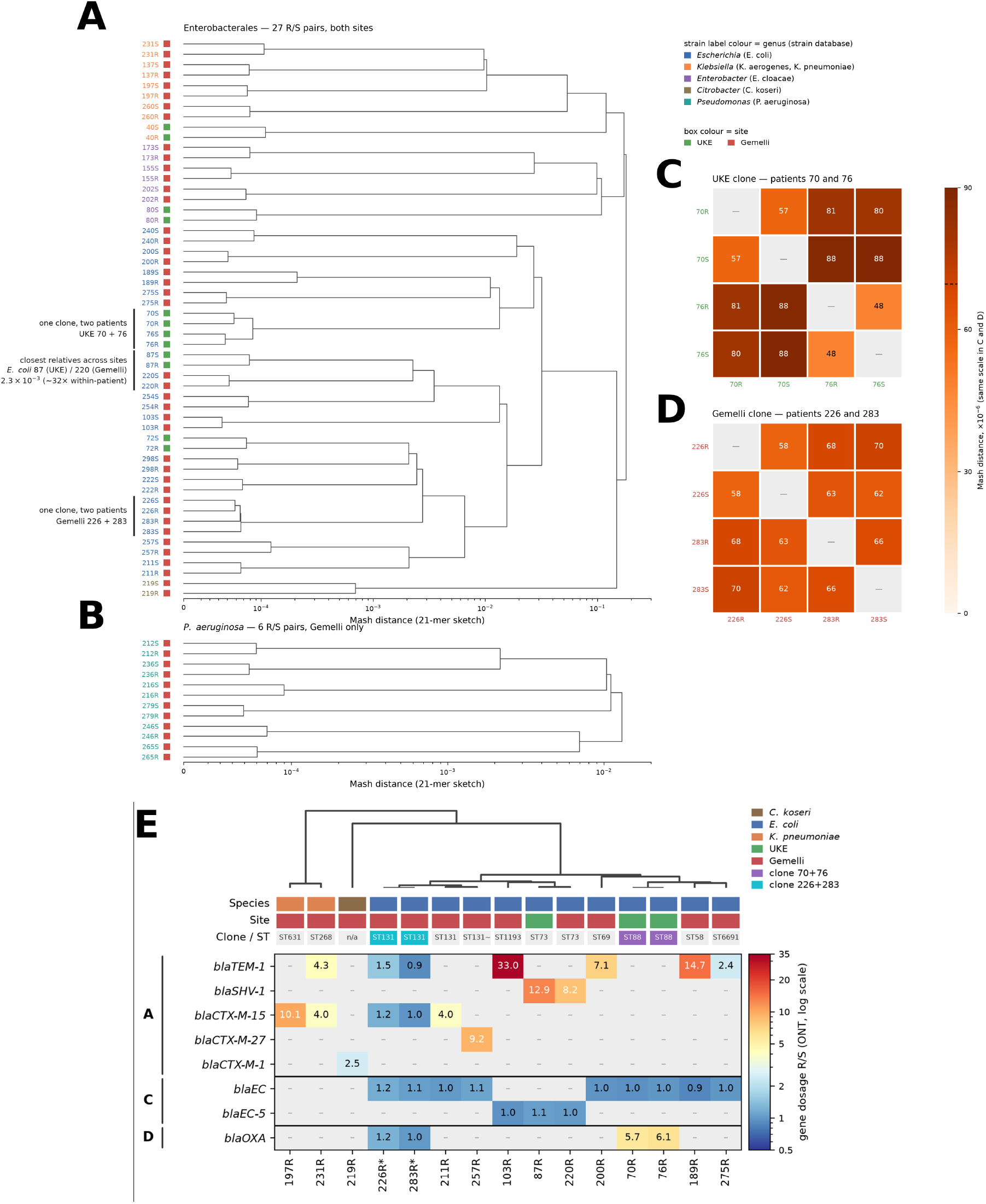
Clonality of the resistant/susceptible pairs. (A) Mash-distance dendrogram of the 27 Enterobacterales resistant/susceptible pairs from both sites; label colour denotes genus and the adjacent box denotes site (green, UKE; red, FPG). (B) The same for the six P. aeruginosa pairs, FPG only. (C, D) Pairwise Mash distances (x10^-6, common scale) within the two within-clone clusters sampled from two patients each - UKE patients 70 and 76 (C) and FPG patients 226 and 283 (D). (E) Long-read (ONT) gene dosage of beta-lactamase genes in the resistant member relative to its paired susceptible member, for the 13 confirmed amplifications plus two sub-threshold pairs (asterisks); colour and printed value give the R/S dosage ratio on a log scale, a dash means the gene is absent from the assembly and 1.0 means equal dosage in both members. The bracketed letters A, C and D to the left of the rows are Ambler classes (A, ESBL/penicillinase; C, AmpC; D, OXA), not panel labels. Column annotations give species, site and clone/sequence type; the tree above is a Mash-type distance on canonical 21-mers (UPGMA), not a core-gene phylogeny. Isolate 211R carries two chromosomal blaCTX-M-15 copies (4.0x and 0.9x) and the amplified copy is shown. Every pair meets the clonality criterion of at least 99.99% average nucleotide identity. FPG is labelled Gemelli in the panel E legend.

A candidate mechanism was resolved in 18 pairs. In 13 (table 1) the resistant member carried a tandem amplification spanning a beta-lactamase gene, with read-depth copy-number ratios from 2.4x to 33.0x relative to the paired susceptible assembly and amplified units of 5-136 kb. The amplification was plasmid-borne in 10 of 13 pairs and chromosomal in 3, and the amplified determinant was *bla*TEM-1 in 5 pairs, *bla*CTX-M-15 in 2, *bla*SHV-1 in 2, *bla*OXA-1 in 2, and *bla*CTX-M-1 and *bla*CTX-M-27 in one each; one *K. pneumoniae* pair co-amplified *bla*TEM-1 and *bla*CTX-M-15 within a single 41 kb unit. The amplified units carried little beyond the beta-lactamase and its mobile-element context (figure 2C, 2D): most contained an integron-associated cassette (*aadA1, qacE*delta1, *sul1*) or plasmid replication and transfer genes, and none contained an additional acquired carbapenemase.

**Table 1:** Resistant/susceptible pairs with tandem amplification of a beta-lactamase locus. Copy-number ratio is read-depth at the amplified gene in the resistant member divided by the paired susceptible member, mapped to the least-duplicated assembly of the pair. MIC is that of the bulk population.

| Pair | Species | Replicon | Amplified resistance gene(s) | Amplified unit (kb) | Copy-number ratio R/S at gene | MIC, bulk (mg/L) | PAP AUC | log10 surviving fraction at 4x BP |
| --- | --- | --- | --- | --- | --- | --- | --- | --- |
| 70 | *E. coli* | chromosome | *blaOXA-1* | 22.0 | 5.7 | 8 | 22.17 | -0.94 |
| 76 | *E. coli* | chromosome | *blaOXA-1* | 21.0 | 6.1 | 8 | 23.13 | -4.78 |
| 87 | *E. coli* | plasmid | *blaSHV-1* | 117.8 | 12.9 | <=4 | 24.7 | -1.51 |
| 103 | *E. coli* | plasmid | *blaTEM-1* | 9.0 | 33.0 | <=4 | 22.24 | -1.6 |
| 189 | *E. coli* | plasmid | *blaTEM-1* | 102.0 | 14.7 | <=4 | 22.44 | -6.43 |
| 197 | *K. pneumoniae* | plasmid | *blaCTX-M-15* | 8.0 | 10.1 | <=4 | 20.06 | -6.28 |
| 200 | *E. coli* | plasmid | *blaTEM-1* | 136.1 | 7.1 | <=4 | 17.58 | -4.98 |
| 211 | *E. coli* | chromosome | *blaCTX-M-15* | 51.0 | 4.0 | <=4 | 24.78 | -0.79 |
| 219 | *C. koseri* | plasmid | *blaCTX-M-1* | 5.0 | 2.5 | <=4 | 22.11 | -2.32 |
| 220 | *E. coli* | plasmid | *blaSHV-1* | 11.0 | 8.2 | <=4 | 18.88 | -5.0 |
| 231 | *K. pneumoniae* | plasmid | *blaTEM-1*; *blaCTX-M-15* | 41.0 | 4.3; 4.0 | 8 | 22.63 | -2.24 |
| 257 | *E. coli* | plasmid | *blaCTX-M-27* | 22.0 | 9.2 | <=4 | 16.74 | -4.09 |
| 275 | *E. coli* | plasmid | *blaTEM-1* | 108.7 | 2.4 | <=4 | 25.37 | -0.75 |

Two *E. coli* pairs from UKE (pairs 70 and 76) were the same ST88 clone sampled two weeks apart and showed an increase in *bla*OXA-1 module dosage between the two sampling points, with amplicon copies at 100% nucleotide identity - the signature of ongoing, recent amplification rather than a fixed duplication. The two isolates recovered from patient 70 differed by 57 x 10-^6^ in Mash distance and those from patient 76 by 48 x 10^-6^, while every cross-patient comparison within the clone fell between 80 and 88 x 10^-6^ (figure 3C): the four genomes therefore share more than 99.99% average nucleotide identity, and the within-patient pairs are marginally closer to each other than to the second patient’s pair, as expected if a single circulating lineage was independently sampled twice. Against that near-identical background the *blaOXA-1* module carried 5.7 times the dosage of its susceptible counterpart in the first pair and 6.1 times in the second (figure 3E, table 1). A second, temporally resolved instance comes from FPG, where the *E. coli* ST131 pairs 226 and 283 are two sampling points, two weeks apart, of a single persistent bloodstream infection in one patient (figure 3D). All four genomes lie within 58 to 70 x 10^-6^ of one another - the distance between the two time points (62 to 70 x 10^-6^) is indistinguishable from the distance between the resistant and susceptible member sampled on the same day (58 and 66 x 10^-6) - so two weeks of persistent bacteraemia under therapy produced no measurable diversification of the core genome. What changed was dosage of *blaTEM-1, blaCTX-M-15, blaEC* and *blaOXA-1*, but below the threshold in a not significant way (figure 3E). The most informative comparison is between sites. *E. coli* 87 from UKE and *E. coli* 220 from FPG were the closest relatives across the two centres, yet still 2.3 x 10^-3^ apart - roughly 32 times the within-patient distances above, or about 99.8% identity, which places them in the same sequence type (ST73) but excludes recent transmission between Hamburg and Rome. Both nevertheless amplified the same gene, *blaSHV-1*, to comparable dosage (12.9-fold and 8.2-fold; figure 3E, table 1). Two epidemiologically unlinked isolates converging on the same amplified determinant is evidence that the amplification is selected by the exposure rather than inherited as part of a locally successful clone, which is the practically relevant point: this phenotype cannot be anticipated from lineage surveillance and will recur wherever the selective pressure and a permissive repeat architecture coincide.

In the remaining 5 pairs (table 2) no amplification was detectable and the resistant member differed by point variants in the *AmpC* regulatory circuit or in efflux machinery. Three were fixed in the resistant population: a nonsense change truncating *ampD* at codon 95 of 188 in the *K. aerogenes* piperacillin case (pair 40), a missense change at the *ampR* effector pocket (D135A) in an *E. cloacae* pair (pair 80), and an *ampD* A97V change at a buried, fully conserved site in a second *E. cloacae* pair (pair 202) - all three predicted to derepress chromosomal *AmpC*, by loss of function of *ampD* or gain of function of *ampR*. Two *P. aeruginosa* pairs carried changes in *dacB* (PBP4), whose inactivation is an established route to *AmpC* hyperproduction: a start-codon loss fixed in pair 216 and a subclonal N312K change (allele fraction 0.60) at a buried residue 8.1 A from the catalytic serine in pair 246. Pair 216 additionally carried two subclonal, tolerated substitutions at the same codon of an RND efflux membrane-fusion protein (allele fractions 0.53 and 0.50), which we do not interpret as causal.

**Table 2:**
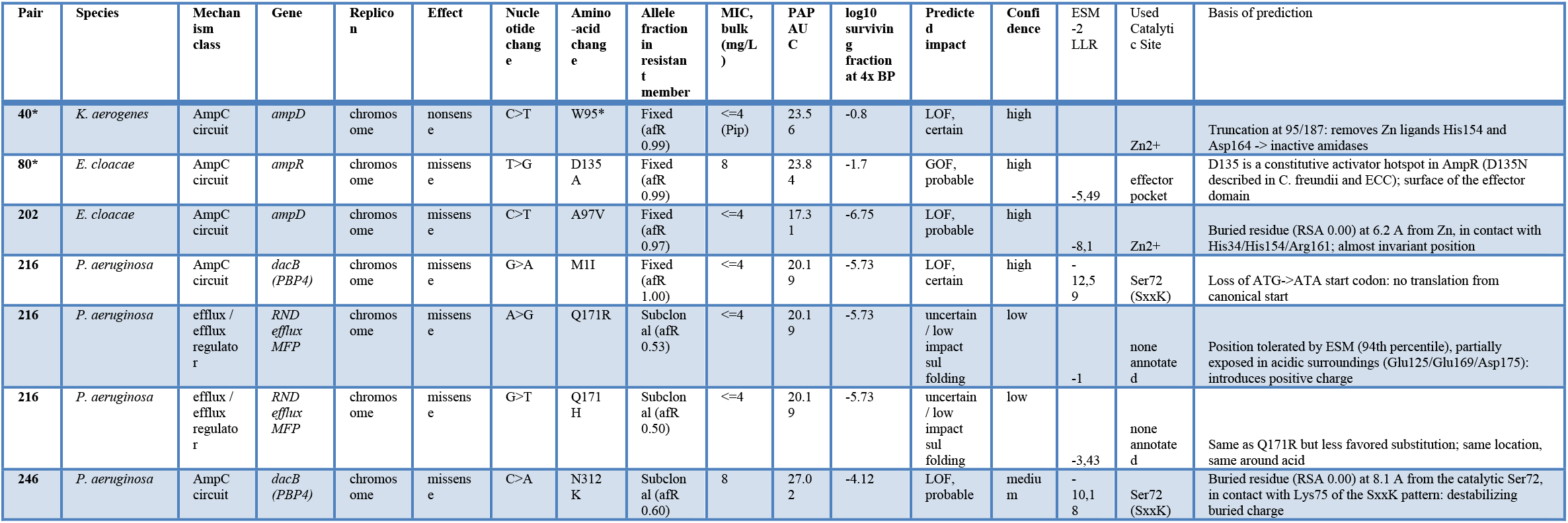
Resistant/susceptible pairs with point variants in the AmpC circuit or efflux machinery. Allele fraction refers to the resistant member. Predicted impact combines the ESM-2 log-likelihood ratio and percentile, relative solvent accessibility, AlphaFold model confidence and distance to the catalytic site; the full annotation, including the rationale for each call, is in the supplementary variant table.

## Discussion

In this prospective, two-centre study we found that heteroresistance to first-line β-lactams is a common, reproducibly detectable phenomenon among Gram-negative bloodstream isolates that routine AST reports as susceptible. In the low AMR burden north Germany center (UKE), 6.45% of tested isolates (95% CI 2.99–13.37) were PAP-confirmed heteroresistant; while in the high AMR burden center Italy center, 14.97% (28/187) of isolates were positive, for a pooled prevalence of 12.1% (34/280). Because every isolate had already been categorised susceptible in the diagnostic laboratory, each of these heteroresistant isolates represents a result that standard-of-care testing would report — and a clinician would act on — as fully treatable, while harbouring a resistant subpopulation able to expand under therapy.Three findings give the phenomenon mechanistic and clinical shape.

First, heteroresistance was almost entirely confined to piperacillin/tazobactam: 33 of 34 HR isolates across both centres were piperacillin/tazobactam-heteroresistant (21.9% of piperacillin/tazobactam-screened isolates), against 1.4% for piperacillin and none for meropenem, and most were high-frequency (surviving fractions ≥10^−2^ across the HR zone). Given how heavily piperacillin/tazobactam is used as empirical Gram-negative cover, this concentration of HR in a single first-line agent is its most clinically consequential feature.

Second, unstable tandem gene amplification was the dominant genomic mechanism, accounting for 11 of 33 evaluable pairs and for the large majority of pairs in which any lesion was found, consistent with the amplification-driven model of Gram-negative HR [1,5]. This mechanism offers a parsimonious explanation for one of the unresolved features of the MERINO trial, in which piperacillin/tazobactam failed to show non-inferiority to meropenem for bloodstream infection caused by ceftriaxone-non-susceptible *E. coli* and *K. pneumoniae* despite *in-vitro* susceptibility of the enrolled isolates [15]. Two observations from that trial sit awkwardly with a simple susceptible-versus-resistant reading of the entry criterion. First, centralised re-testing reclassified a substantial minority of enrolled isolates as non-susceptible to piperacillin/tazobactam, and the discordance was not attributable to a single method or a single site [16]. Second, the isolates that failed were enriched for *blaOXA-1* and *blaCTX-M*, and their MICs rose steeply with inoculum [17,18]. Both features are expected consequences of a copy-number-driven, unstable resistant subpopulation rather than of measurement error. An isolate whose resistant fraction lies near the detection limit of a standard inoculum will be reported susceptible or non-susceptible depending on which cells the inoculum happens to sample, which reproduces the poor between-test agreement observed on re-testing; the same isolate will appear frankly non-susceptible at a higher inoculum, because the resistant subpopulation is then present in absolute numbers sufficient to grow. The genes we found amplified — blaTEM-1, blaCTX-M-15, blaOXA-1 — are exactly those enriched among the MERINO failures, and amplification raises enzyme dosage without altering the hydrolytic spectrum, so it is invisible to genotype-based prediction and reverts on subculture in the absence of drug. We cannot test this hypothesis retrospectively, since population analysis was not performed on the MERINO isolates and stored subcultures would in any case have lost unselected amplifications. The implication is prospective: trials comparing a β-lactam/β-lactamase-inhibitor combination with a carbapenem should characterise the resistant subpopulation, not only the bulk MIC, at enrolment. Amplified units ranged from 3.5x to 36x and were usually multi-gene blocks that bundled a β-lactamase (*bla*TEM-1, *bla*SHV-1, *bla*CTX-M-15, *bla*CTX-M-27 or *bla*OXA-1) with integron-borne aminoglycoside, sulphonamide, tetracycline and quaternary-ammonium determinants and, in several pairs, a *mer* operon: a single amplification event therefore co-amplifies multiple resistance and biocide-tolerance determinants. Notably, one clone was recovered from two different patients at the same centre carrying the identical amplified unit (HR70/HR76), indicating that an amplification-competent lineage can circulate rather than arising *de novo* in each host.

Third, in nearly half the pairs heteroresistance is not written in the DNA sequence at all: 15 of 33 pairs were negative on both genomic arms, and HR72 — negative in two independent sequencing runs — is the best-supported such case, pointing to non-genetic, expression-level heteroresistance [6]. The mechanistic spectrum we observed spans tandem amplification, de-repression of efflux (*ramR, mexR*), candidate PBP4 (*dacB*) lesions predicted to de-repress AmpC transcriptionally, and purely phenotypic cases; no single molecular assay will capture all of it.

A fourth finding concerns measurement rather than biology, and directly constrains how HR should be quantified. Resequencing the six first-centre pairs reproduced every amplification call — the same genes, the same block, the same member — but the measured dosage moved by +20%, −17% and −55% between two runs of the same isolates. Amplicon copy number is therefore a snapshot of subclone frequency in the culture that happened to be sequenced, not a stable strain property. This has two consequences. It retracts the escalation reading we had drawn from the single-run data on the HR70/HR76 clone, where the two isolates in fact carry the same block at comparable dosage. And it sets the specification for the dPCR assays now in development: a single copy-number value is not a strain attribute to be reported once, so assays must be read as quantitative, condition-dependent measurements, ideally alongside the drug exposure and growth history of the sample. A related caution applies to variant-level evidence: in the *P. aeruginosa* subset, 30 of 52 regulator variant calls repeated the identical codon change in independent pairs and were exclusively transitions, the error signature of the long-read chemistry, so cohort-scale recurrence filtering — not per-genome plausibility — was what separated signal from artefact.

The tested resistant subpopulations showed no evident growth defect relative to their bulk populations is clinically important: a low or absent fitness cost means these subpopulations are not efficiently purged when antibiotic pressure is removed, favouring their persistence between exposures and their availability to expand at the next treatment course. Coupled with the demonstration that pre-existing HR can drive within-patient conversion from susceptible to resistant [8], our data support the view that β-lactam HR in GN-BSI is a plausible, under-recognised contributor to empirical-therapy failure.

Our study has limitations. Only two centres contributed, with non-overlapping enrolment periods and different allocation mixes, so the pooled prevalence should not be read as a European estimate; the borderline between-centre difference is confounded by that mix. Allocation to a single investigated agent per isolate means the prevalence for each drug is conditional on the routine phenotype that led to it, and an isolate heteroresistant to more than one beta-lactam would have been detected against only one. The frequency cut-off separating high-from low-frequency heteroresistance is a study convention, as no standardised threshold exists. Genomic characterisation is complete for the pairs reported here but not yet for the full set of confirmed cases, so the relative weight of amplification versus AmpC-circuit variants may shift. Fitness cost of the resistant subpopulations, and the clinical outcome of patients whose isolate was heteroresistant, were not assessed in this analysis; both are the subject of ongoing work. These limitations define the next steps, which we are already pursuing: digital PCR assays personalised to each amplicon (including a *bla*OXA-1-specific assay) to quantify copy number directly and rapidly in clinical material; systematic fitness characterisation of the resistant subpopulations; and, ultimately, a prospective clinical study to test whether breakpoint-crossing HR to first-line β-lactams predicts therapeutic failure in Gram-negative bloodstream infection. Detecting HR is at present laborious and confined to reference laboratories; a validated, rapid molecular readout would be the prerequisite for asking whether acting on HR changes outcomes.

In conclusion, roughly one in eight AST-susceptible Gram-negative bloodstream isolates in this study harboured a heteroresistant subpopulation to a first-line β-lactam — almost always to piperacillin/tazobactam, driven by unstable gene amplification in a third of sequenced pairs and by no detectable genomic lesion in nearly half, and without an obvious fitness penalty. These isolates are invisible to current diagnostics but carry the seed of on-treatment resistance, and they warrant both better detection tools and a direct test of their clinical consequences.

## Data Availability

All data produced in the present study are available upon reasonable request to the authors

https://github.com/damianosquitieri96-hr/hr-heteroresistance-pipeline

## Declaration of generative AI and AI-assisted technologies

During preparation of this work the authors used an AI-assisted scientific computing platform (Claude, Anthropic) for data analysis, and drafting/editing of the text under continuous author supervision All results and figures have been verified against the source data. Responsibility for the content remains with the authors, who declare that they have reviewed and approved the use of the tool in preparing the manuscript.

## Contributors

Conceptualization: DS, HR, MS; Methodology: DS; Investigation: DS; Formal analysis: DS; Data curation: DS; Writing – original draft: DS; Writing – review & editing: DS, GM, BB, NDB, BP, HR, MS; Supervision: HR, MS; Funding acquisition: HR, MS. All authors had full access to the data and had final responsibility for the decision to submit for publication.

